# Curating Retrospective Multimodal and Longitudinal Data for Community Cohorts at Risk for Lung Cancer

**DOI:** 10.1101/2023.11.03.23298020

**Authors:** Thomas Z. Li, Kaiwen Xu, Neil C. Chada, Heidi Chen, Michael Knight, Sanja Antic, Kim L. Sandler, Fabien Maldonado, Bennett A. Landman, Thomas A. Lasko

## Abstract

Large community cohorts are useful for lung cancer research, allowing for the development and validation of predictive models. A robust methodology for (1) identifying lung cancer and pulmonary nodules from electronic health record (EHRs) as well as (2) associating longitudinal data with these conditions is needed to optimally curate cohorts at scale from clinical data. Both objectives present the challenge of labeling noisy multimodal data while minimizing assumptions about the data structure specific to any institution. In this study, we leveraged (1) SNOMED concepts to develop ICD-based decision rules for building a cohort that captured lung cancer and pulmonary nodules and (2) clinical knowledge to define time windows for collecting longitudinal imaging and clinical concepts. We curated three cohorts with clinical concepts and repeated imaging for subjects with pulmonary nodules from our Vanderbilt University Medical Center. Our approach achieved an estimated sensitivity 0.930 (95% CI: [0.879, 0.969]), specificity of 0.996 (95% CI: [0.989, 1.00]), positive predictive value of 0.979 (95% CI: [0.959, 1.000]), and negative predictive value of 0.987 (95% CI: [0.976, 0.994]). for distinguishing lung cancer from subjects with SPNs. This work represents a strategy for high-throughput curation of multi-modal longitudinal cohorts at risk for lung cancer from routinely collected EHRs.

## 1. Introduction

The use of predictive models to inform clinical diagnosis, management, and prognosis is an area of intense research, especially in the early diagnosis of lung cancer from detected pulmonary nodules[1,2]. Large representative cohorts are a key ingredient in developing and validating predictive models that generalize well across communities[3]. Although prospective clinical trials such as the National Lung Screening Trial [4] have provided a richly annotated datasets for this purpose, they are costly to replicate at scale and are limited in scope as they only include high-risk, lung cancer screening patients. Without well-funded clinical trial enrollment, electronic health records (EHRs) represent the next best window into clinical populations[5,6]. Curating a retrospective cohort from the EHRs is a two-step pipeline that includes (1) defining a phenotype to separate cases and controls within an appropriate time window, and (2) mining data across modalities and time.

Individuals with an indeterminate pulmonary nodule (IPN) detected incidentally or during screening, and without a recent or active history of any cancer, represent a clinical challenge due to limitations of available noninvasive methods to risk stratifying IPNs [7]. In contrast, individuals with an active cancer or recent cancer history who present with an IPN undergo are generally managed with more aggressive diagnostic investigations due to a higher pretest probability of malignancy. The value of predictive models is limited in this setting, so these individuals should excluded from study cohorts for lung cancer prediction [8,9]. A common starting point for finding diagnoses from the EHR are International Classification of Diseases (ICD) codes, a hierarchical terminology of medical findings, diagnoses, and conditions that is ubiquitously used for reimbursement requests in the United States[10]. For many diagnoses, including lung cancer, there is no consensus on which ICD codes should be included to define the diagnostic event. Furthermore, identifying cases where an IPN ultimately diagnosed as lung cancer is a nontrivial issue as the information is often only accessible as non-structured data within biopsy reports and clinical notes. This study proposes a strategy for defining lung cancer and IPN events based on existing SNOMED-CT concepts[11]. We further leverage the implicit timing between the two events to label cases and controls.

Once cases and controls have been identified, data from these subjects are commonly retrospectively extracted. An imaging study would require chest CT scans that capture SPNs, ideally repeated scans that show nodule change over time. To this end, imaging studies undertake expensive and time-consuming visual assessments of each image. Studies of non-imaging risk factors likewise undertake challenging efforts to extract clinical concepts from the EHR. These challenges motivate a scalable method for medical image and clinical concept mining that would enable high-throughput research or at least preliminary curation to minimize manual effort. This study proposes to implicitly curate images and clinical concepts that occur in clinically-informed time windows surrounding the lung cancer or SPN events.

Standardized cohort curation methods are needed to increase the chance that cohorts are comparable across geographic and institutional boundaries. However, the underlying data structure of EHRs differ by institution, with each facing unique challenges in extracting information from heterogeneously structured, sparse, and irregularly sampled data. The methods put forth in this study seek to be agnostic to data structure by inferring phenotypes from ICD codes only. We test the validity of these inferences by comparing our cohorts with our institution’s cancer registry [12]. The proposed method was used to curate three cohorts from our home institution: a clinical concepts cohort and two longitudinal imaging cohorts.

## 2. Data

All data were collected from Vanderbilt University Medical Center (VUMC) under a protocol approved by the Vanderbilt Human Research Protections Program, IRB #140274. Non-imaging data were pulled from the Research Derivative, our archive of 2.5 million EHRs from VUMC collected over the two decades [13]. The full history of ICD codes and their occurrence date were retrieved for each subject in the study. We also tapped ImageVU, our linked imaging archive that contains an incomplete subset of chest and full body CTs acquired at VUMC after 2012.

## 3. Methods

Risk factors, biomarkers, and predictive models are most valuable when they inform early risk stratification before patients undergo invasive procedures and well before the disease becomes metastatic. We choose to retrospectively capture this population by finding individuals with a SPN detected incidentally or by screening who do not have a history of any cancer. We use ICD-based rules to define the presence of pulmonary nodules, lung cancer, and history of any cancer, and leverage their relative timing to distinguish those who developed lung cancer from those with benign disease. These methods are used to curate three different cohorts that represent populations from VUMC with (1) an SPN, (2) an SPN and longitudinal chest CT imaging, and (3) just longitudinal chest CT imaging. We denote these cohorts as VU-SPN, VU-LI-SPN, and VU-LI respectively.

### 3.1. ICD-based Phenotypes

ICD-based phenotypes can be inferred using clinical expert-designed schemas that map high level clinical concepts to aggregations of ICD codes. The leading expert-designed schemas that have emerged include Phecodes [14,15], representing diseases for PheWAS-based clinical and genetic research, and SNOMED-CT, a comprehensive terminology that broadly includes clinical concepts beyond diseases. The phenotyping efforts in this study leveraged a mapping between SNOMED-CT concepts and ICD codes [16], but we note that Phecodes result in similar phenotype definitions for lung cancer and pulmonary nodules.

For the SPN phenotype, we used SNOMED-CT with SCTID 427359005, concept name “Solitary nodule of lung (finding)”, to identify ICD-9 793.11 and ICD-10-CM R91.1 both named “solitary pulmonary nodule”. For the lung cancer phenotype, we aggregated the descendants of SCTID 363358000, concept name “Malignant tumor of lung”, and mapped them to ICD-9/ICD-10/ICD-10-CM codes, ultimately finding 56 matching codes in our archives (Table 1). This aggregation of codes represents a broad phenotype of lung cancer and includes any malignancy found in the bronchus or lung, but excludes malignancies of the trachea, larynx, mediastinum, and pleura. The phenotype can be further factorized to distinguish between primary lung cancer and metastasis to the lung from other cancers if the need arises. Finally, a phenotype for any malignancy was created by aggregating the descendants of SCTID 363346000, concept name “Malignant neoplastic disease” and mapping the concepts to ICD codes.

**Table 1.** ICD-based phenotypes for SPN and lung cancer.

| Version | Code | Description |
| --- | --- | --- |
| **Phenotype: Solitary Pulmonary Nodule** |  |  |
| ICD-9 | 793.11 | Solitary pulmonary nodule |
| ICD-10 | R91.1 | Solitary pulmonary nodule |
| **Phenotype: Lung cancer** |  |  |
| ICD-9 | 162† | Malignant neoplasm of trachea bronchus and lung |
| ICD-9 | 197.0 | Secondary malignant neoplasm of lung |
| ICD-9 | 209.21 | Malignant carcinoid tumor of the bronchus and lung |
| ICD-9 | 176.4 | Kaposi's sarcoma, lung |
| ICD-10 | C34* | Malignant neoplasm of bronchus and lung |
| ICD-10 | C7A.090 | Malignant carcinoid tumor of the bronchus and lung |
| ICD-10 | C46.5* | Kaposi's sarcoma of lung |
| ICD-10 | C78.0* | Secondary malignant neoplasm of lung |

†Includes all sub-categories below the hierarchy except 162.0 “Malignant neoplasm of trachea”. \*Includes all sub-categories below the hierarchy under this general category.

### 3.2 Criteria for inclusion, case, and control

We defined the cohort inclusion criteria as individuals with a SPN phenotype and no cancer phenotype occurring before the SPN phenotype (Figure 1). Lung cancer cases are individuals with a lung cancer phenotype occurring 4 to 1095 days after the SPN phenotype. A SPN that is stable for three years is highly unlikely to be malignant, based on previous clinical studies. Controls are individuals that meet the inclusion criteria but not the positive case criteria. Importantly, we excluded records that ended within three years of an SPN. We defined the end of a record as the date of the last ICD code plus a 1 month buffer. These rules were used to label VU-SPN and VU-LI-SPN.

**Figure 1.**
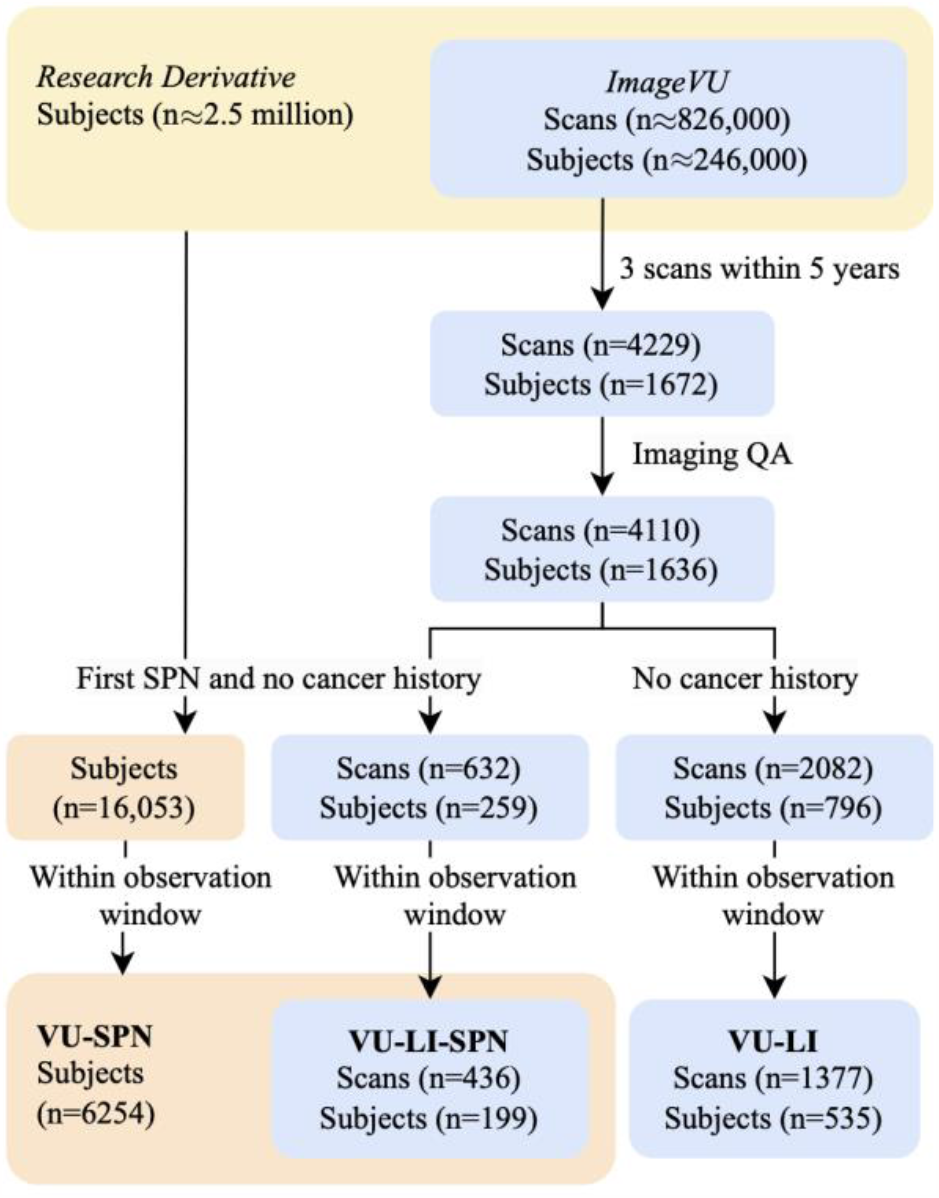
Archives linking EHRs to imaging allowed for the selection of subjects via ICD rules. Scans that were low quality and data that did not fall within observation windows were excluded. VU-SPN: subjects with no cancer history prior to an SPN code. VU-LI-SPN: subjects in VU-SPN with imaging. VU-LI-Incidence: subjects with imaging.

These criteria represent a conservative strategy that may not be adequately sensitive for capturing lung cancer incidence, since subjects must have a SPN that rises to the threshold of being worked up to be included in the cohort. We defined a broader inclusion criteria to identify those with and without lung cancer, regardless of SPN presence. Cases were those without cancer of any type before an occurrence of a lung cancer phenotype. Controls were those without lung cancer, and no cancer of any type before an observation. Any data occurring after a diagnosis of cancer were excluded. These rules were used to label VU-LI.

### 3.3 SPN cohort

We collected records from the Research Derivative with ICD codes matching the SPN phenotype. Our observation window for each subject ranged inclusively from the start of their record to the date of their lung cancer event. Within this window, we collected demographics, ICD codes, laboratory values, and medication orders. Observations occurring after the lung cancer code was excluded (Table 2).

**Table 2.** Cohorts Characteristics.

| **Cohort** | **VU-SPN** | **VU-LI-SPN** | **VU-LI** |
| --- | --- | --- | --- |
| No. subjects | 6254 | 199 | 535 |
| Cases/Controls | 946 (6%) / 5308 | 30 (15%) / 169 | 66 (12%) / 469 |
| No. scans | N/A | 436 | 1337 |
| Cases/Controls | N/A | 42 (9.9%) / 394 | 88 (6.6%) / 1249 |
| Age | 57.2±15.8 | 59.9 ±13.1 | 62.0 ±11.0 |
| Sex (male) | 2776 (44%) | 126 (59%) | 383 (72%) |
| BMI | 29.2±7.03 | 27.5±7.23 | 27.1±6.33 |

### 3.4 Longitudinal Imaging cohorts

We assembled a cohort with repeated chest CTs that captured pulmonary nodules or untreated lung cancer for a longitudinal imaging study (Figure 1). We started with an initial discovery cohort of individuals in ImageVU with three CTs within five years. As a quality assurance step, we algorithmically analyzed the imaging metadata to discard images with poor slice contiguity and unrealistic physical dimensions. We also performed a fast manual review to remove CTs that did not fully include the lung field or had occluding artifact. Finally, we retrieved ICD codes for the discovery cohort that passed this quality assurance and identified cases and controls (Table 2).

A unique challenge in building imaging cohorts is inferring which images best capture a lung cancer without the need for visual assessment or robust natural language processing of radiologic reports. For lung cancer cases we implicitly classified images based on their timing relative to the first occurring lung cancer event (Figure 2). The classes are distinguished as follows. *Pre-3+*: Images acquired three or more years before the lung cancer phenotype. They are unlikely to capture any relevant pulmonary nodules. *Pre-3*: Images acquired 1-3 years before the lung cancer phenotype. They are likely to capture pulmonary nodules in the pre-malignant stage. *Pre-1*: Images acquired from the date of the lung cancer phenotype to 1 year before. They are likely to capture undiagnosed and untreated lung cancer [8,9]. *Post-3*: Images acquired 3 years after the lung cancer phenotype was observed. They are likely to capture lung cancer that was diagnosed and treated. *Post-3+*: Images acquired more than 3 years after the lung cancer phenotype. They are not likely to capture findings relevant to lung cancer. For controls, we designate two classes of images as useful for analysis: images before the SPN code (*Pre*) and those within three years after the SPN (*Post-3*). Images acquired more than three years after the SPN (*Post-3+*) were discarded due to the possibility of containing unlabeled lung cancer.

**Figure 2.**
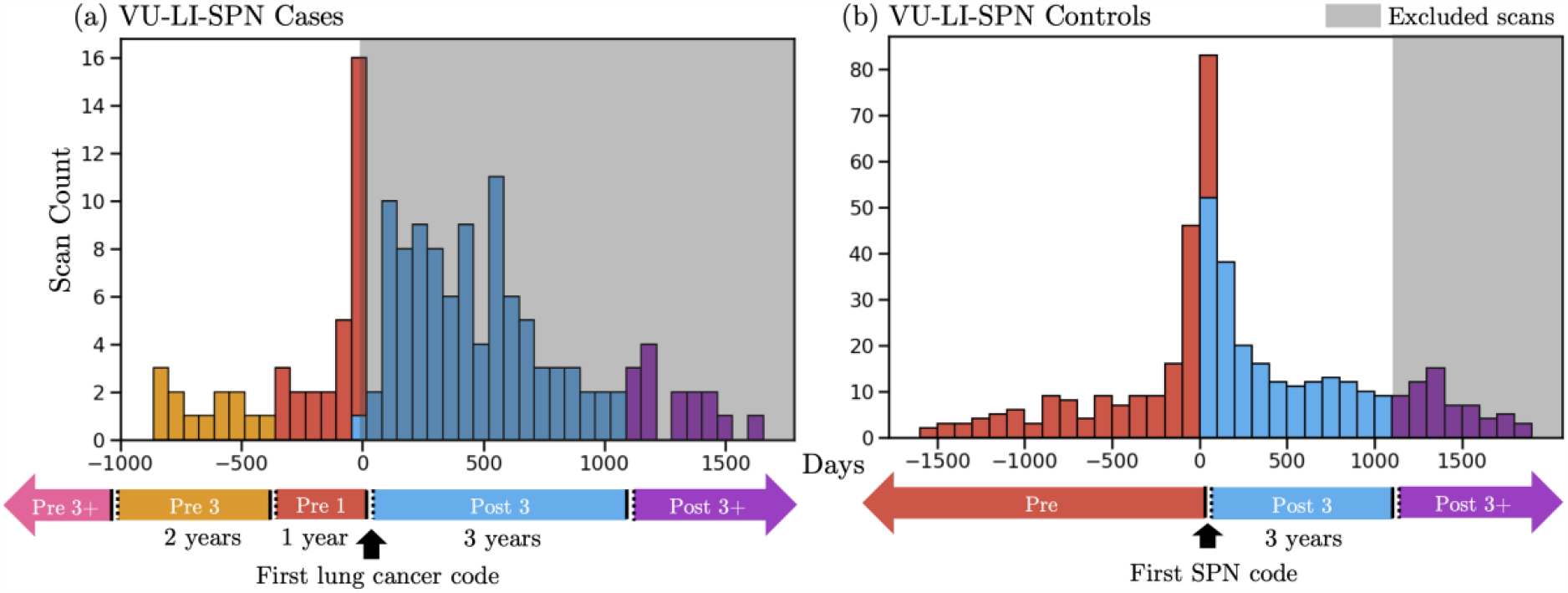
Distribution of scans surrounding first diagnosis of lung cancer in the LIVU-SPN cohort. Scans were classified into disjoint time windows (in chronological order: Pre 3+, Pre 3, Pre 1, Post 3, and Post 3+) based on their proximity to the first lung cancer event for cases or first SPN event for controls. For cases (a), scans occurring at or before the lung cancer event (Pre 3+, Pre 3, Pre 1) were included in the cohort while scans collected after were excluded (Post 3, Post 3+). For controls (b), scans that were acquired before or within three years after the first SPN code (Pre, Post 3) were included in the cohort while scans acquired three years after were excluded (Post 3+).

### 3.5 Validation

The ICD-based decision rules for distinguishing lung cancer cases and controls were compared against the VUMC Cancer Registry (VCR), an externally developed registry of all patients who received a cancer diagnosis or first course treatment for a cancer at VUMC from 1983 to 2023. For inclusion in the registry, records are first broadly selected using pathology reports or the presence of ICD codes. Each selected record is reviewed by trained clinicians and confirmed cases are reported the Tennessee State Registry. We estimate that this process produces an extremely low false positive rate for inclusion in the VCR to indicate a true cancer case [12]. However, the false negative rate is difficult to bound because the VCR does not include patients diagnosed at other institutions who then receive second course treatment or beyond at VUMC.

To explain the gap between our cohorts and the VCR, we conducted a chart review of the mismatched patients using clinical notes and pathology reports (Table 3). Due to the large cohort size, we reviewed a random 10% of cases and controls absent from the VCR. We did not review cases present in the VCR because they are manually reviewed and we expected a negligible false positive rate.

**Table 3.**
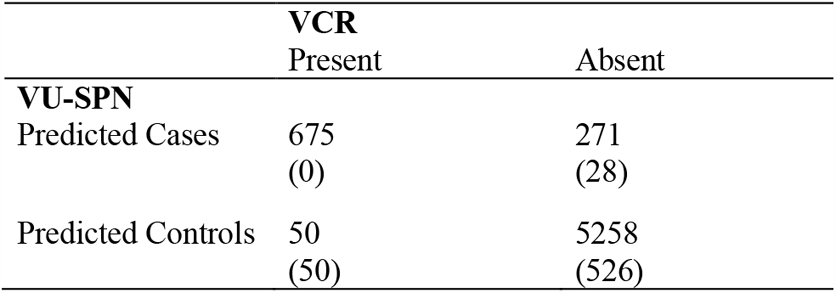
VU-SPN cases/controls vs. presence in VUMC Cancer Registry (VCR). (Number of subjects that we chart reviewed from each cell.)

|  | VCR |  |
| --- | --- | --- |
|  | Present | Absent |
| **VU-SPN** |  |  |
| Predicted Cases | 675<br>(0) | 271<br>(28) |
| Predicted Controls | 50<br>(50) | 5258<br>(526) |

### 3.6 Statistics

We used the following bootstrap procedure to estimate the proportions of cases and controls that truly meet criteria from our chart review. First, we attained 100,000 samples by sampling with replacement from subjects whose charts were reviewed. The size of each sample was 627, which is 10% of VU-SPN. We stratified the sampling by the comparison between VU-SPN and VCR. That is, each bootstrapped sample was the union of a 10% sample from the 675 cases present in VCR, a 100% sample from the 28 reviewed cases absent from VCR, a 10% sample from the 50 reviewed controls present in VCR, and a 100% of the 526 reviewed controls absent from VCR. We report the proportion estimates as the bootstrapped medians. Values at the 2.5^th^ and 97.5^th^ percentile among bootstrap samples formed the 95% confidence intervals of each estimate (Table 4). We also computed the sensitivity, specificity, positive predictive value (PPV), and negative predictive value (NPV) in each bootstrap sample and report their aggregate estimates using the same procedure.

**Table 4.** Estimated proportion of predicted cases and controls in VU-SPN that truly met criteria, reported as median and 95% CI of bootstrapped samples.

|  | Estimated True Case | True Control | Do not meet inclusion criteria |
| --- | --- | --- | --- |
| **VU-SPN** |  |  |  |
| Predicted Cases | 0.979<br>[0.948, 1.00] | 0.021<br>[0.00, 0.052] | 0<br>[0, 0] |
| Predicted Controls | 0.013<br>[0.006, 0.024] | 0.987<br>[0.976, 0.994] | 0.009<br>[0.002, 0.019] |

For imaging cohorts, we simply conducted reviewed the predicted cases absent from VCR and predicted controls present in the VCR (Table 5). We did not perform a full review of these imaging cohorts because we conducted our validation with a larger overlapping cohort in VU-SPN.

**Table 5.**
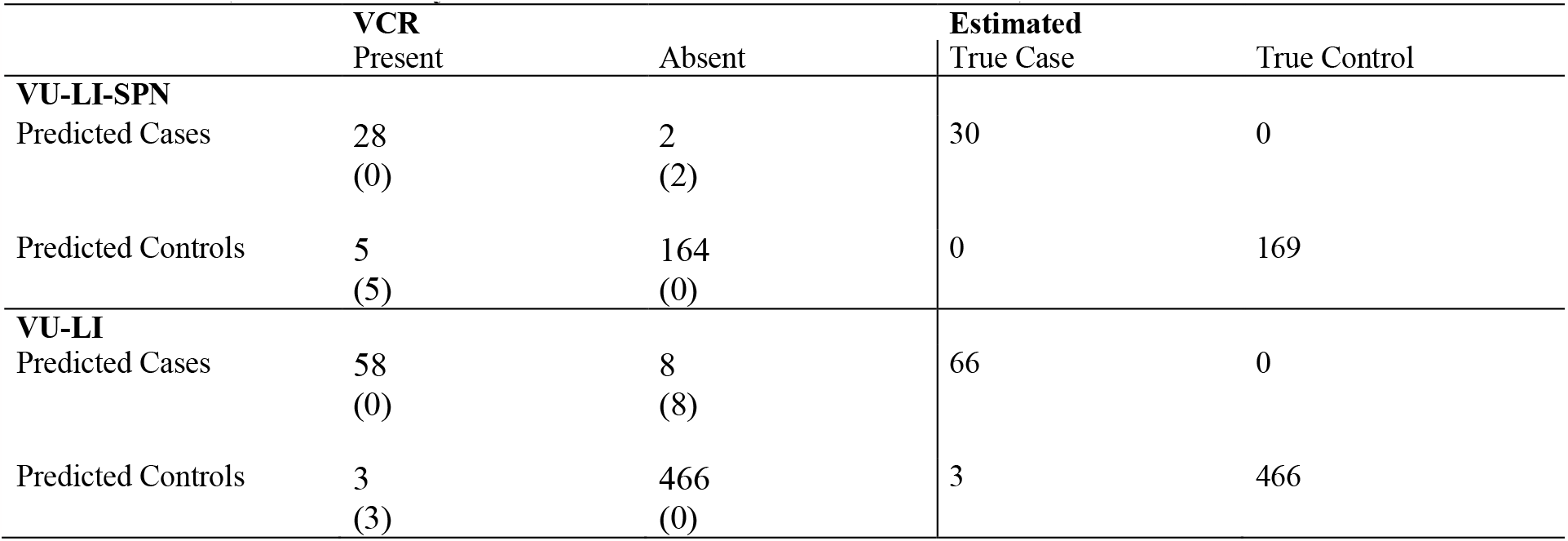
Estimated true cases and controls from VU-LI-SPN and VU-LI. Only mismatches between cohort vs. VCR were reviewed. (Number of subjects that we chart reviewed from each cell)

|  | VCR |  | Estimated |  |
| --- | --- | --- | --- | --- |
|  | Present | Absent | True Case | True Control |
| **VU-LI-SPN** |  |  |  |  |
| Predicted Cases | 28<br>(0) | 2<br>(2) | 30 | 0 |
| Predicted Controls | 5<br>(5) | 164<br>(0) | 0 | 169 |
| **VU-LI** |  |  |  |  |
| Predicted Cases | 58<br>(0) | 8<br>(8) | 66 | 0 |
| Predicted Controls | 3<br>(3) | 466<br>(0) | 3 | 466 |

## 4. Results

### 4.1 Clinical Concepts

16,053 unique subjects were found to match inclusion criteria. However, 9769 controls were excluded due to their record ending within three years of the SPN date. Ultimately we identified 946 cases and 5308 controls (Table 2). We collected all demographics, ICD codes, laboratory tests, and medications occurring before the SPN.

### 4.2 Longitudinal Imaging

4229 CT scans across 1672 subjects were included in the initial discovery cohort. From the discovery cohort, 4110 chest CTs across 1636 subjects were found to meet quality standards. 199 of these subjects met the SPN inclusion criteria with 30 lung cancer cases and 169 controls. The broader inclusion criteria identified 535 subjects with 66 cases and 469 controls.

VU-LI-SPN cases were associated with 167 chest CTs with 0 in the Pre-3+ class, 13 in Pre-3, 29 in Pre-1, 94 in Post-3, and 31 in Post-3+ (Figure 2a). Controls were associated with 465 chest CTs with 189 in the Pre class, 205 in Post-3, and 71 in Post-3+ (Figure 2b). VU-LI cases were associated with 2082 chest CTs, with 1 in the Pre-3+ class, 16 in the Pre-3 class, 71 in the Pre-1 class, 543 in Post-3, and 202 in Post-3+. Since images in the Post-3 and Post-3+ class are likely to capture cancers that have been diagnosed and treated, their diagnostic value to an imaging study is uncertain and they should excluded. After excluding usable scans, VU-LI-SPN captured 436 scans across 199 subjects while the VU-LI captured 1337 scans across 535 subjects (Table 2).

### 4.3 VCR Validation

In the VU-SPN cohort we reviewed all 50 controls present in the VCR, 28 out of 271 cases absent from the VCR, and 451 out of 5258 controls absent from the VCR. Within the first group, 4 (8%) were diagnosed with lung cancer before the SPN date, 10 (20%) were diagnosed within three years after the SPN, and 36 (72%) were diagnosed beyond three years after the SPN. Within the second group, we found that 24 records met case criteria while 4 were unable to be confirmed as cases via chart review. 2 of these 4 subjects were likely to have lung cancer based on the clinical picture, but the diagnosis was not confirmed due to patient choice and patient death. For the third group, we found 1 (0.19%) subject with lung cancer, 5 (0.95%) subjects with a history of cancer before their SPN, and 520 (98.8%) subjects that met control criteria. With bootstrapping, we estimated that 0.979 (95% CI: [0.948, 1.00]) of predicted cases and 0.987 (95% CI: [0.976, 0.994]) of predicted controls to truly meet their respective criteria (Table 4). Our method achieved a median sensitivity of 0.930 (95% CI: [0.879, 0.969]), specificity of 0.996 (95% CI: [0.989, 1.00]), and positive predictive value of 0.979 (95% CI: [0.959, 1.000]), negative predictive value of 0.987 (95% CI: [0.976, 0.994]).

In the VU-LI-SPN cohort, there were 5 controls present in the VCR and 2 cases absent from the VCR. All of the former developed lung cancer more than three years after their first observed SPN code, meaning they were appropriately labeled as a control. Chart review of the latter confirmed that they all met case criteria despite being absent from the VCR. In VU-LI there were 3 controls present in the VCR and 8 cases absent from the VCR. Chart review determined that all of the former did have lung cancer, while all of the latter met case criteria (Table 5).

## 5. Discussion

In this work we outline a strategy that leverages simple and well-defined rules around ICD codes to curate three cohorts for studying pulmonary nodules at risk for lung cancer from our local institution. Our approach avoids any systematic assumptions about the institution or the EHR, except for similarity in the use of the relevant ICD codes for reimbursement purposes. Within these cohorts we verify that our approach is highly accurate in identifying subjects with and at risk for lung cancer. We are not surprised that lung cancer codes have high specificity, at 0.996, and high PPV, at 0.979, because billing for this life-changing condition should not occur unless clinicians are certain of the diagnosis. We believe this is a reasonable explanation for our results that likely holds across code sets of other cancers and across different institutions.

In this work, we excluded a large portion of data because it fell outside of the observation windows of interest. The observation window for non-imaging data was anytime before the SPN event, while the window for imaging data depended on its proximity to the lung cancer and SPN events. This strategy is suitable for building a validation cohort because it prevents estimates of the posterior probability, found in data after the lung cancer event, from leaking into the validation. However, including data that occurs after the lung cancer event can be beneficial for hypothesis generation or model development, as this research may gain insight from seeing posterior observations. For example, unsupervised training on imaging acquired after diagnosis of lung cancer can lend statistical strength to a predictive model even if those images have no diagnostic value.

A few edge cases demonstrate the limitations of our approach. First, there were 4 controls in VU-LI-SPN that became lung cancer cases in VU-LI. These were subjects that had a lung cancer code which occurred three years after their first SPN code. The lung cancers are most likely unrelated to the first SPN and may have arisen from other nodules that the subjects acquired after the first. The inability to distinguish between multiple nodules in the same subject is a limitation of our approach.

Second, our validation supports a 7% false negative rate with various modes of failure. 14 out of the 20 false negatives developed lung cancer but were incorrectly billed and did not receive a lung cancer code. 5 of the false negatives were subjects who had a clinical note citing a remote history of cancer before their SPN and therefore should not have met our inclusion criteria. There was no corresponding ICD code for these subjects. A single false negative had a code for mucosa-associated lymphoid tissue lymphoma (MALT), which can arise in the lung and present as a SPN [17]. However, ICD taxonomy does not distinguish pulmonary MALT lymphoma from MALT lymphoma in other organs. In summary, our high-throughput method is effective at curating and labeling cohorts for lung cancer research from subjects that have a EHR footprint in the form of billing codes, but rare limitations arise when relying on the medical billing system.

## Data Availability

All data produced in the present study are available upon reasonable request to the authors

## References

[1] F.S. Collins, H. Varmus, A New Initiative on Precision Medicine, New England Journal of Medicine. 372 (2015) 793–795. 10.1056/NEJMP1500523/SUPPL_FILE/NEJMP1500523_DISCLOSURES.PDF.

[2] E.P. Gray, M.D. Teare, J. Stevens, R. Archer, Risk Prediction Models for Lung Cancer: A Systematic Review, Clin Lung Cancer. 17 (2016) 95–106. 10.1016/J.CLLC.2015.11.007.

[3] A. Halevy, P. Norvig, F. Pereira, The unreasonable effectiveness of data, IEEE Intell Syst. 24 (2009) 8–12. 10.1109/MIS.2009.36.

[4] Reduced Lung-Cancer Mortality with Low-Dose Computed Tomographic Screening, New England Journal of Medicine. 365 (2011) 395–409. 10.1056/NEJMOA1102873/SUPPL_FILE/NEJMOA1102873_DISCLOSURES.PDF.

[5] M.R. Cowie, J.I. Blomster, L.H. Curtis, S. Duclaux, I. Ford, F. Fritz, S. Goldman, S. Janmohamed, J. Kreuzer, M. Leenay, A. Michel, S. Ong, J.P. Pell, M.R. Southworth, W.G. Stough, M. Thoenes, F. Zannad, A. Zalewski, Electronic health records to facilitate clinical research, Clin Res Cardiol. 106 (2017). 10.1007/S00392-016-1025-6.

[6] M.S. Lauer, R.B. D’Agostino, The Randomized Registry Trial — The Next Disruptive Technology in Clinical Research?, New England Journal of Medicine. 369 (2013) 1579–1581. 10.1056/NEJMP1310102/SUPPL_FILE/NEJMP1310102_DISCLOSURES.PDF.

[7] P.P. Massion, R.C. Walker, Indeterminate Pulmonary Nodules: Risk for Having or for Developing Lung Cancer?, Cancer Prevention Research. 7 (2014) 1173–1178. 10.1158/1940-6207.CAPR-14-0364.

[8] H. MacMahon, D.P. Naidich, J.M. Goo, K.S. Lee, A.N.C. Leung, J.R. Mayo, A.C. Mehta, Y. Ohno, C.A. Powell, M. Prokop, G.D. Rubin, C.M. Schaefer-Prokop, W.D. Travis, P.E. Van Schil, A.A. Bankier, Guidelines for management of incidental pulmonary nodules detected on CT images: From the Fleischner Society 2017, Radiology. 284 (2017) 228–243. 10.1148/RADIOL.2017161659/ASSET/IMAGES/LARGE/RADIOL.2017161659.FIG14B.JPEG.

[9] M.P. Rivera, A.C. Mehta, M.M. Wahidi, Establishing the Diagnosis of Lung Cancer: Diagnosis and Management of Lung Cancer, 3rd ed: American College of Chest Physicians Evidence-Based Clinical Practice Guidelines, Chest. 143 (2013) e142S–e165S. 10.1378/CHEST.12-2353.

[10] W.H. Organization, International statistical classification of diseases and related health problems 10th revision., (2011).

[11] C. Gaudet-Blavignac, V. Foufi, M. Bjelogrlic, C. Lovis, Use of the Systematized Nomenclature of Medicine Clinical Terms (SNOMED CT) for Processing Free Text in Health Care: Systematic Scoping Review, J Med Internet Res 2021;23(1):E24594 Https://Www.Jmir.Org/2021/1/E24594. x23 (2021) e24594. 10.2196/24594.

[12] M.L. Riyad Naser, Judith Roberts, Todd Salter, Jeremy L Warner, An informatics-enabled approach for detection of new tumor registry cases, J Registry Manag. 41 (2014) 19–23.

[13] I. Danciu, J.D. Cowan, M. Basford, X. Wang, A. Saip, S. Osgood, J. Shirey-Rice, J. Kirby, P.A. Harris, Secondary Use of Clinical Data: the Vanderbilt Approach, J Biomed Inform. 52 (2014) 28. 10.1016/J.JBI.2014.02.003.

[14] W.Q. Wei, L.A. Bastarache, R.J. Carroll, J.E. Marlo, T.J. Osterman, E.R. Gamazon, N.J. Cox, D.M. Roden, J.C. Denny, Evaluating phecodes, clinical classification software, and ICD-9-CM codes for phenome-wide association studies in the electronic health record, PLoS One. 12 (2017) e0175508. 10.1371/JOURNAL.PONE.0175508.

[15] P. Wu, A. Gifford, X. Meng, X. Li, H. Campbell, T. Varley, J. Zhao, R. Carroll, L. Bastarache, J.C. Denny, E. Theodoratou, W.Q. Wei, Mapping ICD-10 and ICD-10-CM Codes to Phecodes: Workflow Development and Initial Evaluation, JMIR Med Inform. 7 (2019). 10.2196/14325.

[16] National Institutes of Health, SNOMED CT to ICD-10-CM map, U.S. National Library of Medicine. (n.d.). https://www.nlm.nih.gov/research/umls/mapping_projects/snomedct_to_icd10cm.html.

[17] R. Borie, M. Wislez, M. Antoine, C. Copie-Bergman, C. Thieblemont, J. Cadranel, Pulmonary mucosa-associated lymphoid tissue lymphoma revisited, European Respiratory Journal. 47 (2016) 1244–1260. 10.1183/13993003.01701-2015.

